# Multiple Imputation Approaches for Missing Time-to-Event Outcomes with Informative Censoring: Practical Considerations from a Simulation Study Based on Real Data

**DOI:** 10.1101/2024.11.18.24317244

**Authors:** Andrea Bellavia, Min Guo, Sabina A. Murphy

**Affiliations:** TIMI Study Group, Division of Cardiovascular Medicine, Brigham and Women’s Hospital, Harvard Medical School, Boston, MA, United States; Department of Biostatistics, Harvard T.H. Chan School of Public Health, Boston, MA, United States

**Keywords:** missing data, multiple imputations, survival analysis, informative censoring

## Abstract

Missing outcomes data represent a common threat to the validity and robustness of clinical trials and prospective epidemiologic studies with time-to-event outcomes. Several studies have outlined the importance of critically evaluating missing outcome data in clinical studies, as well as the relevance of multiple imputations (MI) in this context. Recent MI extensions, namely controlled-MI, have been introduced as a viable alternative for sensitivity analysis in the presence of informative censoring, yet they lack validation based on real data. In this study we used data from a randomized trial to generate realistic scenarios of potential censoring mechanisms, used to assess the practical relevance of several imputation approaches for missing outcome data. Our results confirm the relevance of multiple imputations approaches, especially in studies with long follow-up and higher proportion of potentially informative censoring. This first study comparing MI and controlled-MI approaches for missing outcome data can help practitioners appreciate the advantages of different imputation approaches under realistic settings in prospective studies in clinical epidemiology.

## Main text

Missing outcomes data represent a common threat to the validity and robustness of clinical trials and prospective epidemiologic studies with time-to-event outcomes. The most common reason for outcome missingness in these contexts is censoring, which generally occurs when patients are lost to follow-up or withdraw consent from an ongoing clinical study. Several studies have outlined the importance of critically evaluating missing outcome data in clinical studies, as well as the relevance of multiple imputations (MI) in this context, and researchers are expected to justify their handling of missing data. ^1–4^ Statistical approaches commonly used to report primary results in prospective studies, such as the non-parametric Kaplan-Meier estimator or the Cox regression model, generally provide valid estimates of the effect of interest under the assumption that censoring mechanisms are unrelated to either exposure or outcome. This assumption, commonly referred to as non-informative censoring, corresponds to an assumption of missingness at random for missing outcome data. There are several settings, however, where this assumption might not be met. In a clinical trial, for example, occurrence of adverse or safety events could lead to drug discontinuation and potential study withdrawal. If the occurrence of a given adverse event is more likely to occur for a specific treatment regime, this would lead to a higher risk of censoring in a specific study arm (i.e. informative censoring). Recent MI extensions, namely controlled-MI, have been introduced as a viable alternative for sensitivity analysis in the presence of informative censoring. ^5^ In particular, γ-imputation provides a flexible tool that allows incorporating existing knowledge on the potential censoring mechanisms to evaluate how the treatment effect would change under different scenarios. ^6^ In brief, γ-imputation relaxes the non-informative censoring assumption of the Cox model by including a γ term, corresponding to the log-HR comparing censored and uncensored individuals, and the uses MI to impute censored observations based on this modified Cox model. ^7^

Existing literature and recommendations for missing outcome data are largely based on theoretical considerations and simulation studies alone. Simulations are often conducted based on general assumptions for the censoring distributions and study population, and limited information from real data is available for clinical researchers and epidemiologists. To such end, in this study we used data from the TIMI-58 DECLARE trial, ^8^ a randomized, double-blind, multinational, placebo-controlled, phase 3 trial, to generate realistic scenarios of potential censoring mechanisms based on real data. This large clinical study (n=17,160) experienced very limited censoring due to withdraw consent or loss to follow-up (<2%), and we therefore evaluated it as the real target population. The detailed information on the occurrence of safety events and drug discontinuation collected was used to generate fictitious alternative settings of censoring that are likely to occur in real settings. First, we evaluated a potential scenario where all adverse events that led to drug discontinuation (affecting ∼7.5% of participants) also led to trial follow-up discontinuation and therefore censoring. As several of these adverse events are associated with treatment, such censoring would be informative. Next, under the same censoring mechanisms and trial characteristics, we generated additional fictitious trials gradually increasing the proportion of censored individuals, the length of follow-up, and the proportion of trial participants experiencing the primary event. We then applied MI and γ-based controlled-MI to impute censored observations under all settings, comparing bootstrapped HRs from imputed data to the HRs from the complete data reported in the study. When using controlled-MI, γ values are provided by the user. In our example we used reported information on adverse events to define individual-specific γ values as the inverse of the log(HR) reported in the trial safety results. In practice, when the effect of the main exposure (e.g. treatment) on censoring is unknown, it is recommended to evaluate a range of potential γ values based on clinical background and a-priori hypothesis.

Table 1 summarizes results from the different evaluated settings. Results indicate that both standard MI and controlled-MI provide robust inference under all evaluated scenarios, with slightly improved performances of controlled-MI only with heavy censoring (∼30%) and longer follow-up (10 years). These results confirm the relevance of multiple imputations approaches for missing outcome data in prospective studies. We join previous researchers in recommending their inclusion as sensitivity analyses in study protocols and statistical analysis plans, especially in studies with long follow-up and higher proportion of potentially informative censoring. Controlled-MI, and in particular γ-imputation, can be used to assess the extent to which potential informative censoring would affect primary trial results, thus providing a powerful tool for sensitivity analysis for prospective studies in the presence of missing outcome data and censoring for which reasons are unknown.

**Table 1.**
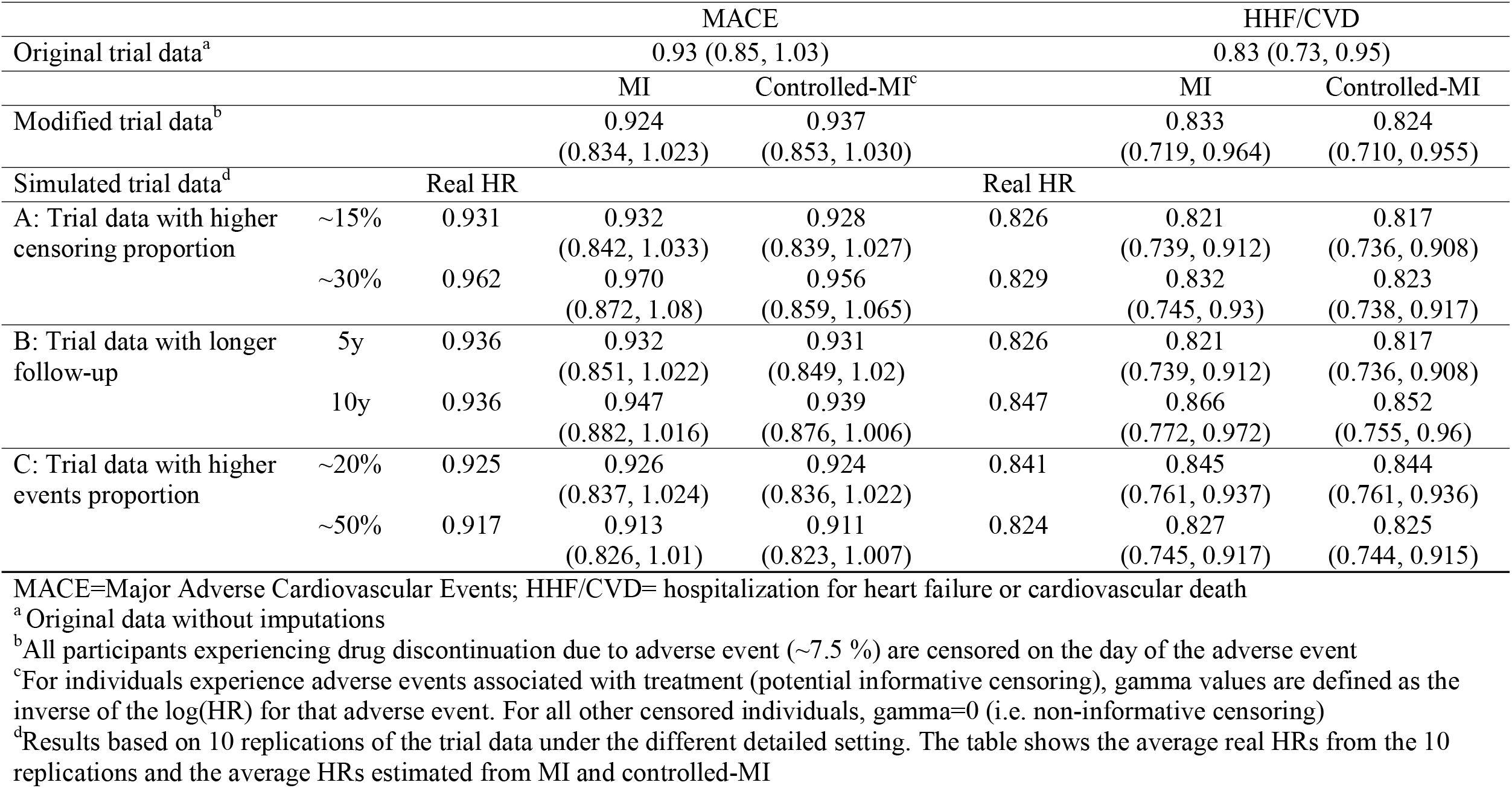
Estimates of treatment effects (dapagliflozin vs placebo) for primary endpoints of DECLARE TIMI-58 using multiple imputations and controlled multiple imputations, under different replications of the trial data summarizing settings with informative censoring.

To our knowledge, this is the first study comparing MI and controlled-MI approaches for missing outcome data using extensive simulated scenarios that are based on real data and censoring mechanisms occurring in applied clinical and epidemiologic research. By generating fictitious replications of a complete clinical study and using observed information on adverse events mechanisms to generate censoring, this study can help practitioners appreciate the advantages of different imputation approaches under realistic settings in prospective studies in clinical epidemiology.

## Data Availability

All data produced in the present study are available upon reasonable request to the authors

## References

1. Bell, M. L., Fiero, M., Horton, N. J. & Hsu, C.-H. Handling missing data in RCTs; a review of the top medical journals. BMC Med. Res. Methodol. 14, 118 (2014).

2. National Research Council (US) Panel on Handling Missing Data in Clinical Trials. The Prevention and Treatment of Missing Data in Clinical Trials. (National Academies Press (US), Washington (DC), 2010).

3. Groenwold, R. H. H., Donders, A. R. T., Roes, K. C. B., Harrell, F. E., Jr & Moons, K. G. M. Dealing With Missing Outcome Data in Randomized Trials and Observational Studies. Am. J. Epidemiol. 175, 210–217 (2012).

4. Altman, D. G. Missing outcomes in randomized trials: addressing the dilemma. Open Med. 3, e51–e53 (2009).

5. Tan, P.-T., Cro, S., Van Vogt, E., Szigeti, M. & Cornelius, V. R. A review of the use of controlled multiple imputation in randomised controlled trials with missing outcome data. BMC Med. Res. Methodol. 21, 72 (2021).

6. Jackson, D. et al. Relaxing the independent censoring assumption in the Cox proportional hazards model using multiple imputation. Stat. Med. 33, 4681–4694 (2014).

7. Cro, S., Morris, T. P., Kenward, M. G. & Carpenter, J. R. Sensitivity analysis for clinical trials with missing continuous outcome data using controlled multiple imputation: A practical guide. Stat. Med. 39, 2815–2842 (2020).

8. Wiviott, S. D. et al. Dapagliflozin and cardiovascular outcomes in type 2 diabetes. N. Engl. J. Med. 380, 347–357 (2019).

